# Investigating Informal Sector Workers Preferred Health Insurance Design Features: A Pivotal Position for Engagement in Health Purchasing Decisions in Ghana Using the Q-sort Methodology: A Research Protocol

**DOI:** 10.1101/2025.06.08.25329231

**Authors:** Jemima Sumboh, Helen Elsey, Noemia Siqueira, Marc Suhrcke

**Affiliations:** Department of Health Science, Hull York Medical School, University of York, York, England, United Kingdom; Centre of Health Economics, University of York, York, England

**Author notes:** Current Address: Luxembourg Institute of Socio-economic Research (LISER), Luxembourg (Marc Suhrcke). These authors contributed equally to this work (Helen Elsey, Noemia Siqueira, Marc Suhrcke).

## Abstract

Globally, health insurance has been recognized as a pragmatic health financing option to achieve Universal Health Coverage particularly for the poor and vulnerable. However, the implementation of such programs are at times cumbersome, saddled with and reinforcing existing health inequities. Empirical evidence has shown that the uptake of health insurance is low among informal sector workers. To facilitate the uptake of health insurance, it is important to actively engage healthcare consumers and providers in health purchasing decisions. However, there is a paucity of evidence on the mechanisms employed to engage healthcare consumers and providers in healthcare purchasing decision making which reflects their interests and concerns. This research will therefore seek to identify health insurance design features which will enhance enrollment of informal sector workers unto the Ghana National Health Insurance Scheme from the perspective of informal sector workers, health care providers, and health and policy stakeholders.

To facilitate addressing the research objectives highlighted, this study will use a Q-sort methodology. Q-sort methodology combines the strength of qualitative and quantitative methods for the systematic exploration of subjectivity; people’s perspective, opinion, and beliefs. Study participants are expected to rank different statements under each health insurance feature. The attached weights are analysed, and the prominent statements are highlighted, interpreted, with the support of qualitative interviews. The study will be conducted in La Nkwantanang Madina and Ashaiman, two districts of the Greater Accra Region of Ghana. Purposive and snowball sampling will be used to identify the study participants.

The findings of this research supplements existing efforts to improve engagement between communities, key stakeholders within the health and local government systems to improve primary health outcomes. Empirical evidence from this research will reinforce advocacy for and the formulation of policies specific to improving informal sector workers uptake and use of health insurance.

## 1.1 Introduction

Globally, there is a consensus for member states of the World Health Organization (WHO) to develop their health financing systems in a manner that ensures all people have access to the needed health care services, while being protected against the financial risks associated with using these health services (1). This goal defined as Universal Health Coverage (UHC) requires governments to prioritize and address three key questions when enacting health financing reforms; how is a health system to be financed? How can individuals be protected from the financial risks of ill-health? How can they encourage the optimum use of available resources (1)? Globally, health insurance has been recognized as a pragmatic health financing option to achieve UHC particularly for the poor and vulnerable. However, the implementation of such programs are at times cumbersome, saddled with and reinforcing existing health inequalities or inequities (2,3). Empirical evidence has shown that the uptake of health insurance is low amongst the poor (2,3). Moreover, it is difficult to achieve high coverage among the informal sector (2–5). This is extremely problematic for the urban poor who often work in the informal sector (6), are most burdened by communicable and non-communicable diseases (7), work and reside in areas with a growing health market dominated by private and informal providers (8), and low penetration of health insurance in the private and informal health market (9).

In Ghana, there are three types of health insurance schemes: National Health Insurance Scheme (NHIS), private commercial health insurance schemes, and private mutual health insurance schemes (10). The NHIS was implemented in 2004 and has a comprehensive benefit package which covers about 90 percent of disease conditions in the country. Some of the health care services covered under the NHIS include outpatient services, including diagnostic testing and operations such as hernia repair; inpatient services including specialist care, most surgeries and hospital accommodation, oral health treatments, all maternity care services (antenatal, deliveries (normal, assisted and cesarean) and post-natal), all emergency care, eye care services and all drugs on the National Health Insurance Authority Medicines list (11–13). The NHIS is mandatory for all Ghanaians, unless alternative private health insurance can be demonstrated. However, in practice, membership is voluntary for the informal sector (10). While formal sector workers contribute directly to the NHIS through a 2.5% deduction of their Social Security and National Insurance Trust (SSNIT) contributions, informal sector workers are required to pay annual premiums based on their income to the NHIS either by visiting the district NHIS or through mobile money payments (13). In reality, however, informal sector workers are charged a flat premium and varies from district to district due to the difficulty of categorizing people into different socio-economic groups (10). The aged (people above 70 years), children under 18 years whose parents are enrolled, indigents, and all pregnant women are exempted from paying premiums (12). Thus, over 60% of active members of the NHIS are under the premium exemption category (13). Members can access services from accredited providers (public, private and mission) and are expected to co-pay for certain health services. The NHIS reimburses providers based on Diagnostic Related Groups and fee-for-service (14). About 41% of the population are currently enrolled unto the NHIS (15). The NHIS has been prioritized as the gateway to achieving Universal Health Coverage. The NHIS is intended to address the challenges in providing and accessing health care services especially to vulnerable populations by reducing financial barriers to accessing health care services and enhancing citizens choice of healthcare providers, amongst others (16,17). Since the inception of the NHIS in 2004, for instance, considerable achievements in financial access to health services have been attained. However, geographical access, equity, quality of care and efficiency in the use of health resources issues remain a challenge (18).

## 1.2 Empirical Evidence of the Existing Challenges of Health Insurance for Informal Sector Workers

The challenges associated with implementing health insurance programs for informal sector workers ranged from the difficulty in encouraging and enforcing enrollments, the difficulty in financially sustaining such health insurance programs or schemes, inequitable contributions to the scheme, and the general poor governance and transparency mechanisms in the management of these schemes. In Ghana, about 59% of the population are without any health insurance cover, with a high percentage being in the informal sector (19,20). This trend is similar to the enrollment rates of the informal sector in Sub-Saharan Africa such as Nigeria, Tanzania and Kenya. In Nigeria, for instance, about 4.5 million people representing 3% of the population are enrolled unto the NHIS, and majority of these enrollees are in the formal sector (21,22). The voluntary nature of the Acts establishing these schemes for the informal sector makes it extremely difficult for governments to enforce mandatory enrollment for informal sector workers (23,24). Besides, in countries such as Ghana and Rwanda, although it is mandatory for members to have a health insurance cover, it is difficult to enforce mandatory enrollment because of the large proportion of the population in the informal sector and weak operational capacities of health insurance purchasing organizations (10).

Also, most people in the informal sector have not enrolled in any health insurance scheme because of high premium rates (non-affordability of the premium) (25), poor trust in the health insurance schemes, long and complicated registration processes (25,26), preferential treatment of patients paying out-of-pocket for health services over members of health insurance schemes by health providers (4), and perceived poor quality of health (27). In Ghana, most private providers prefer private mutual health insurance scheme members to NHIS members to which informal sector workers belong therefore limiting their access to a wide range of healthcare services (27).

Financial unsustainability has also been identified as a challenge of health insurance schemes for informal sector workers. Often, pooled funds for informal sector workers are inadequate and dependent on unsustainable financing sources. Moreso, such pools are susceptible to adverse selection and moral hazard (28,29). In Rwanda for instance, the community-based health insurance (CBHI) financial deficit has risen from 3,896 RWF million in 2011/2012 to 17,670 RWF million in 2017/2018 (30). In Ghana, total claims payment rose from GHS 7.6 million in 2005, to over GHS 1.07 billion in 2015 (31), and the deficit currently GHS 300 million (31). Although the main reasons accounting for the NHIS in Ghana operating at a deficit are tied to the low levels of enrollment and the majority of members of the NHIS belonging to the exempt category and therefore do not pay annual premiums (19), in Nigeria, the concerns over the financial sustainability of the NHIS are linked to the small and fragmented nature of pools which are largely dependent on government and donor subsidies (32).

Another key challenge impending the extension of health insurance to informal sector workers is the regressive nature of contributions to these schemes as these payments are not linked to individuals or households ability-to-pay. Most countries often pay a flat monthly or flat yearly rate. Often, deciding on an income-based rate that will be progressive is challenging especially for informal sector workers who have irregular incomes, and who are in a sector that is inadequately organized (33). Financing of these schemes is therefore not linked to ability-to-pay and payments are often regressive in nature. Moreso in Nigeria, the flat-rate monthly payments made to the Universal Students’ Health Insurance Program (USSHIP) vary depending on the health package chosen. (34). Thus, the poor and vulnerable are more likely to have a limited benefit package and subsequently limited access to accredited health services compared to the wealthy. Additionally, contributors to these schemes may be required to co-pay for certain health services at the point of service use. Such co-payments are most likely regressive in nature since they are often flat rate co-payments or user fees (28). In Ghana, although informal sector workers are supposed to be charged annual premiums based on their income levels such that the lower income group pays about USD1 and the highest income groups pays USD4.14, in practice, a flat rate is charged due to the difficulty in categorising people into different socio-economic groups (10,35).

The way health insurance schemes are governed, and healthcare services are purchased can influence improvements in service delivery processes, cost containment, organisational efficiency, and accountability. From the literature, organizational weaknesses, poor communication strategies, cumbersome accreditation processes, delays in reimbursement of health facilities, poor accountability, or monitoring mechanisms, and a “top-down” approach to decision making have weakened health insurance schemes (36)^-^(37). The top-down approach to decision making and the exclusion of health providers and state governments in decision making results in its low uptake by private health facilities and states (36).

In Ghana, most health facilities complained about the top-down approach to decision making. Often, reimbursement payment rates, prices on the national medicines list and other technical decisions are made without due consultation^-^(37). Also, health providers reported long delays in reimbursement of claims (37).. Some of the reasons listed to account for these delays include lack of pooled funds, inadequate staff to process claims reporting resulting in errors on claims submitted, and lack of feedback on reasons for delays or rejected claims (37). Other challenges such as unclear vetting procedures by officers at the claims processing centre (CPC) and frequent change of claims forms were reported (37). In a study by Akweongo et al (2021), study participants lamented that it could take a year or two without a facility receiving any reimbursement of claims from the NHIA. Another study by Sieverding et al (2018) reported that several respondents found it was burdensome to obtain the papers or copies of other licenses and certificates required to apply for accreditation (38). Additionally, the fact that NHIS has not made provision to pay for preventive and promotive health services rendered under the Ghana Community-based Health Planning and Services (CHPS) Strategy is undermining Ghana’s Primary Health Care (PHC) system. This is because sending clinical nurses to Community-Based Health Planning and Services (CHPS) zones will shift the focus of this system of delivering door-to-door preventive health care to the traditional health facility-based curative health care (39).

## 1.3 Research Problem/Gaps

Although numerous studies have highlighted the low coverage of informal sector workers by health insurance schemes and the impacts of low levels of enrollments on the sustainability of health insurance schemes and the progress towards UHC, very few studies have explored in-depth different mechanisms to enhance enrollments amongst informal sector workers. Often in the literature, researchers recommend the expansion of health insurance schemes to cover the informal sector workers. However, there are no clearly cut plans of how to ensure informal sector workers are covered by health insurance schemes. Also, to facilitate the uptake of health insurance, it is important to actively engage health care consumers (individuals/households) and healthcare providers in health purchasing decisions. However, there is a paucity of evidence on the mechanisms employed to engage healthcare consumers and providers in healthcare purchasing decision making which reflects their interests and concerns. This research will therefore seek to fill in these research gaps by providing new knowledge on a health insurance model, which reflect the preferences, values, and needs of informal sector populations in Ghana. Specifically, the study will seek to identify health insurance design features which will enhance enrollment of informal sector unto the Ghana National Health Insurance Scheme, mechanisms to link informal sector workers ability-to-pay to income-based premiums, and mechanisms to improve the operational and financial sustainability of the Ghana National Health Insurance Scheme for informal sector workers.

## 1.4 General Objective

To identify and examine health insurance design features which have the potential to improve the uptake and use of health insurance amongst informal sector workers in Ghana

## 1.5 Specific Objectives

1. To identify different health insurance design features that have been implemented in Low-and Middle Income Countries
2. To identify the potential health insurance design features that would be feasible to deliver within the Ghanaian context
3. To identify the health insurance design features preferences of informal sector workers’ in Ghana

## 1.6 Research Questions

1. What health insurance design features have been implemented in LMICs for informal sector workers?
2. What health insurance design features can be implemented in Ghana for informal sector workers?
3. What health insurance design features are preferred by informal sector workers in Ghana?

## 2. Methodology

### 2.1 Study Design

This study will use a mixed method design which is embedded in the q-sort methodology to identify the health insurance design features that will improve the uptake and use of health insurance in Ghana. The use of the mixed method design facilitates the triangulation of data and ensures that the subjectivity of participants’ perspectives and the context sensitivity of beliefs are preserved in the quantitative analysis (40).

### 2.2 Conceptual Framework

The Kutzin Health Financing Framework, which supports the descriptive analysis of a country’s health financing system (41), will be adapted to conceptualize the different health insurance design features that can be implemented for the informal sector. The framework highlights and describes each of the key health financing functions, their interactions and interdependence. It is therefore useful as a tool for the identification and preliminary assessment of policy options to improve health financing systems (41). The key policy levers of the framework include sources of pooled funds and contribution methods, pooling of health care revenues which involves the accumulation of prepaid health care revenues on behalf of a population, purchasing of health services which involves the transfer of pooled resources to service providers on behalf of the population for which the funds were pooled, provision of services, benefit package and out-of-pocket payments, and regulations and information to improve policy outcomes (41).

**Figure 1:**
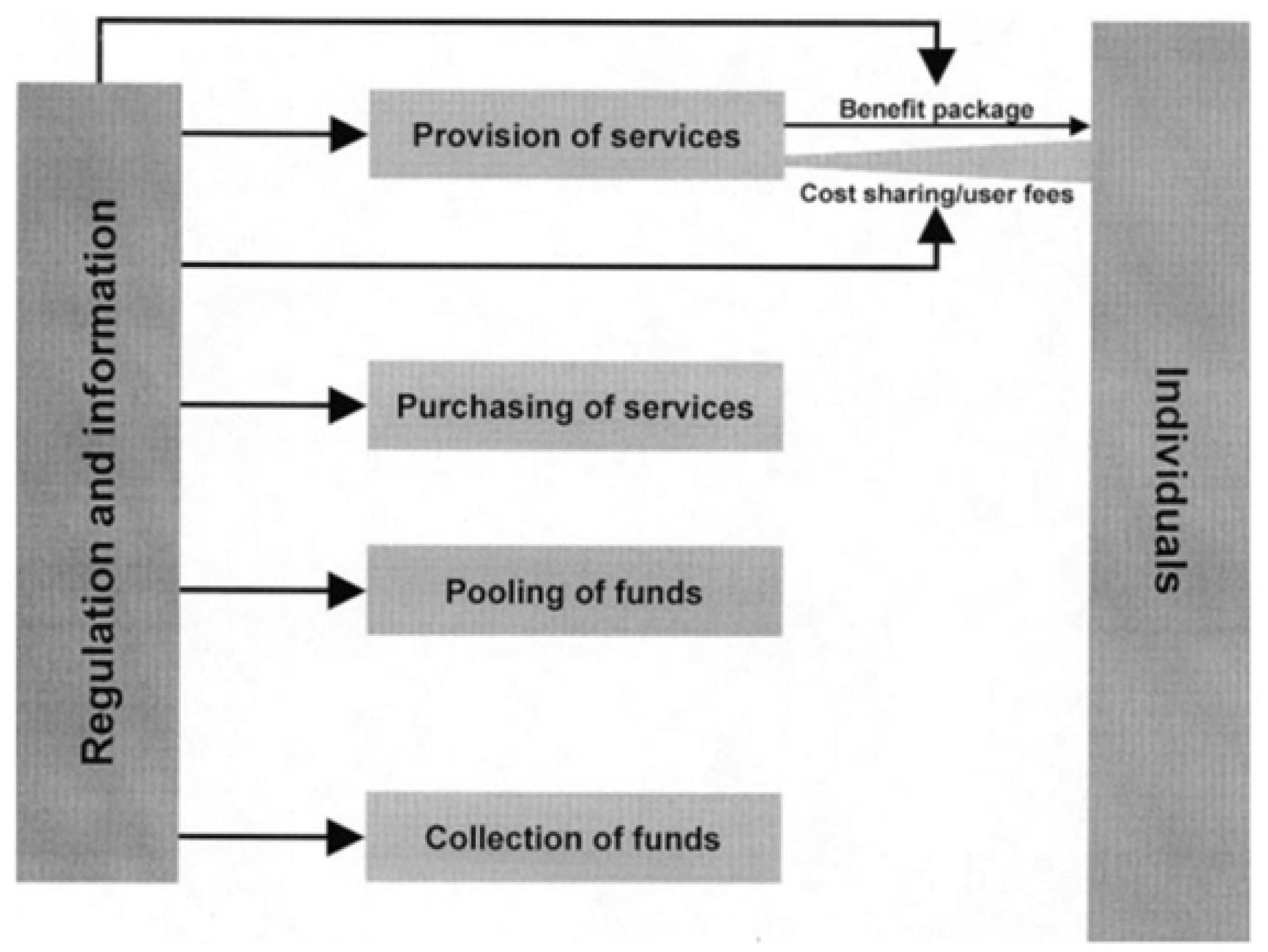
Kutzin (2001) Health Financing Framework.

### 2.3 Specific Research Methods and Justification

In LMICs, different health insurance designs have been implemented to improve population coverage amongst informal sector workers. A pivotal determinant of the success of these health insurance schemes is how to attract and retain informal sector workers on these health insurance plans. However, there are no clearly cut plans of how to ensure informal sector workers are covered by health insurance schemes. As stated above, there is a paucity of evidence on the mechanisms employed to engage healthcare consumers and providers in healthcare purchasing decision making which reflects their interests and concerns. By studying the subjective ranking of healthcare purchasing considerations, this study seeks to identify varied perspectives of consumers.

To facilitate this, this study uses a Q-sort methodology. The Q methodology serves as a foundation for the systematic exploration of subjectivity; people’s perspective, opinion, beliefs, attitude etc. (42–44). It was developed by Stephenson in the 1930s, with its origins in psychology (45), however, it has been applied outside the field of psychology, gaining roots in health system research, with a growing number of published studies spanning a diverse range of health system topics for instance patient perspective of health care, health behaviour and outcomes, and attitudes, beliefs and preferences (46).

This methodology is commonly used to provide new, evolving and potential insights into the study of complex research problems where human subjectivity is a central theme (47). The Q-sort methodology combines the strengths of both qualitative and quantitative methods for the systematic exploration of subjectivity (48). The qualitative component can contribute to the disclosure of subjective perspectives of a defined number of study participants and the quantitative component to ascertain consensus in dealing with distinctive prospects among generally an assertive number of standpoint statements (47). It has therefore been recognized as a qualitative-quantitative hybrid (48).

This methodology enables the exploration of a complex research problem from a participant’s perspective. Basically, study participants are expected to rank different statements based on how they perceive the problem. The patterns that emerge from diverse responses are then used to reveal and label distinct “points of view”, even within small groups (49). Because Q-sort analysis captures the subjectiveness of people’s perspectives, and because the data are easy to gather, analyze and present, Q-methodology is good not only as a research tool but also as a participatory exercise (49).

In this study, the Q-methodology will involve 4 key stages. A detailed description of the q-methodology can be found in (50).

#### 2.3.1 Step 1: Collection and selection of statement sets

The first step in the Q-sort methodology involves the collection of ideas, concepts and statements on the subject in question, in this case, pertaining to health insurance design in health care purchasing, ideally until reaching saturation point (46). The sampling of these statements will be from five main phases. First, relevant statements will be retrieved from an already conducted systematic review that supplements this study. The objective of this systematic review was to evaluate and describe the health insurance mechanisms for informal sector workers, and their impacts on health outcomes and health system goals such as reducing financial risk protection, improving healthcare utilization and quality of health care services amongst informal sector workers in LMICs. The following electronic databases were searched for relevant primary studies; Medical literature Analysis and Retrieval System Online (Medline), Excerpta Medica Database (EMBASE), EconLit Databases via OVIDSP, and Cumulative Index to Nursing and Allied Health Literature (CINAHL), Academic Search Premier and Business Source Premier Databases via EBSCO. The search was limited to empirical studies published between 2011 and September 2022. A total of 391 articles were retrieved from the databases listed above, from which 101 duplicates were removed. The remaining 290 articles were subsequently screened, and 18 articles met the inclusion criteria and were included in this review. Eight studies were quantitative studies employing quasi experimental designs and 10 were qualitative studies. The statements will be retrieved from the findings of the systematic review and specifically based on the characteristics of the health insurance schemes that were retrieved and reported in the review. Table 1 represents the q-sort sampled statements retrieved from the systematic review.

**Table 1:** Q-sort Sampled Statement with Designated Categories.

| Q-sort Sampled Statements with Designated Categories |  |  |
| --- | --- | --- |
| Numb<br>er | Statement | Category |
| 1 | Contribution towards health insurance should be made by individuals | Revenue Collection |
| 2 | Families or households should be charged as a unit | Revenue Collection |
| 3 | Contributions should be made by informal sector workers groups | Revenue Collection |
| 4 | Health insurance should be funded through direct taxes | Contribution Methods |
| 5 | Health insurance should be funded through indirect taxes | Contribution Methods |
| 6 | Contributions should be deducted from microfinance banks informal sector workers belong to | Contribution Methods |
| 7 | Health insurance should be funded through payroll taxes | Contribution Methods |
| 8 | Health insurance should be funded through government transfers | Contribution Methods |
| 9 | Compulsory monthly payments to health insurance scheme | Contribution<br>Methods |
| 10 | Compulsory annual payments to health insurance scheme | Contribution<br>Methods |
| 11 | Voluntary monthly payments to health insurance scheme | Contribution<br>Methods |
| 12 | Voluntary annual payments to health insurance scheme | Contribution<br>Methods |
| 13 | Funds should be pooled by an autonomous/semi-autonomous government purchasing agency | Pooling<br>Organization |
| 14 | Funds should be pooled by a microfinance provider | Pooling<br>Organization |
| 15 | Funds should be pooled by a private insurance company | Pooling<br>Organization |
| 16 | Funds should be pooled by a non-governmental organization | Pooling<br>Organization |
| 17 | Payments should be made based on a percent of salary or income of informal sector workers | Allocation Mechanism |
| 18 | A flat rate payments should be made by informal sector workers | Allocation Mechanism |
| 19 | Different flat rate payments for different health insurance package | Allocation Mechanism |
| 20 | Health services should be purchased by an autonomous/semi-autonomous government purchasing agency | Purchasing Organization |
| 21 | Health services should be purchased by a microfinance provider | Purchasing Organization |
| 22 | Health services should be purchased by a private insurance company | Purchasing Organization |
| 23 | Health services should be purchased by a non-governmental organization | Purchasing Organization |
| 24 | Health services should be purchased using capitation | Provider Payment Methods |
| 25 | Health services should be purchased using case-based payments | Provider Payment Methods |
| 26 | Health services should be purchased using fee-for-service | Provider Payment Methods |
| 27 | Health services should be purchased using a budget | Provider Payment Methods |
| 28 | Health services should be purchased using capitation and case-based payments | Provider Payment Methods |
| 29 | Health services should be purchased using capitation and fee-for-service | Provider Payment Methods |
| 30 | Health services should be provided by accredited private health facilities | Service Providers |
| 31 | Health services should be provided by public health facilities | Service Providers |
| 32 | Health services should be provided by accredited pharmacies, chemical shops | Service Providers |
| 33 | Health services should be provided by independent providers | Service Providers |
| 34 | We should have access to primary health care providers | Level of Service Providers |
| 35 | We should have access to secondary health care providers | Level of Service Providers |
| 36 | We should have access to tertiary health care providers | Level of Service Providers |
| 37 | We should have access to laboratories | Level of Service Providers |
| 38 | We should have access to pharmacies, drugs/chemical shops | Level of Service Providers |
| 39 | We should have access to ultrasounds | Level of Service Providers |
| 40 | The benefit package should include general doctor consultations | Benefit Package |
| 41 | The benefit package should include specialized doctor consultations | Benefit Package |
| 42 | The benefit package should include basic medicines | Benefit Package |
| 43 | The benefit package should include all prescribed medicines | Benefit Package |
| 44 | The benefit package should include diagnostic tests | Benefit Package |
| 45 | The benefit package should include all inpatient care | Benefit Package |
| 46 | The benefit package should include all outpatient care | Benefit Package |
| 47 | The benefit package should include restricted inpatient care | Benefit Package |
| 48 | The benefit package should include restricted outpatient care | Benefit Package |
| 49 | Informal sector workers should be reimbursed their average daily wage | Benefit Package |
| 50 | No out-of-pocket payments | Out-of-pocket payments |
| 51 | Co-payments pegged at a fixed rate | Out-of-pocket payments |
| 52 | Co-payments pegged as a percentage of the total cost | Out-of-pocket payments |
| 53 |  | Regulations and<br>Information to<br>improve policy<br>outcomes |

The second phase, which supplements the online literature search, involves conducting qualitative interviews with a group of key informants, consisting of health care users (informal sector workers), purchasers (NHIS), health providers and the government. The interviews with the key informants are independent of the statements generated from the systematic review. This method to select statement sets is to ensure that the statements are focused and relevant to informal sector works and the Ghanaian context since the systematic review was mainly focused on LMICs. Purposive sampling will be used to identify these key informants. The qualitative interviews will be centred on eliciting responses from the key informants on the health insurance design features which they recommend will improve the uptake and use of health insurance amongst informal sector workers. **S1 Appendix**, **S2 Appendix** and **S3 Appendix** represent the interview guides that will be used in this phase.

In the third phase, the sampled statements will be reviewed by experts in health financing and urban health in Ghana, the Ghana CHORUS team and leaders from informal sector groups, to ensure that the sampled statements have content validity. Content validity is defined as the ability of the selected items to reflect the variables of the construct in the measure (51).

Independent reviews will be conducted to determine which sampled statements are appropriate, accurate and interpretable. The sampled statements are either accepted, rejected or modified based on simple majority opinion (52). The theoretical framework will also be used to enhance content validity.

In the fourth phase, the face validity of the sampled statements (study instrument) will be assessed through a pilot study with research participants. Research participants will be given the opportunity to comment on the difficulty level of understanding the statements, wording, desired suitability and relationship between items and the main objective of the instrument, ambiguity and misinterpretation of items and or comprehensibility (51). This will help reduce systematic and communication measurement errors.

In the final phase, the revised sampled statements are then edited where necessary, reduced and refined (i.e. grouping similar themes together) to produce a manageable Q-set (46). These statements are then randomly assigned numbers.

#### 2.3.2 Step 2: Selection of Population

In q sort methodology, only a limited number of study participants is required, enough to establish the existence of a factor for purposes of comparing one factor with another (42). It involves a structured sample of respondents who are relevant to the research objective. The number of study participants associated with a factor is of less importance relative to who they are (42). The q-sort uses an inverted factor analysis and unlike quantitative surveys, is not focused on generalisability. Studies often have sample sizes of about 40-60 participants (46).

In this study, purposive and snowballing sampling will be used to identify a wide range of study informal sector workers. Purposive sampling will be used to identify the initial study participants from different subgroups of informal sector workers to improve sample diversity. Based on the ICLS classification, informal sector workers will be identified from the following subgroups: self employed workers (e.g. street vendors, home-based workers, farmers and agricultural workers), casual or daily wage workers (e.g. construction and agricultural laborers, and domestic workers), contract or subcontract workers (e.g. industrial outworkers and service sector workers), migrant workers (e.g. seasonal migrants, urban migrants), informal entrepreneurs (e.g. shop owners, service providers, transport operators), unpaid family workers, informal wage workers (e.g. factory, restaurant and hotel staff, manual laborers), domestic and care workers (e.g. housekeepers and cleaners, childcare providers and elderly caregivers) and informal professionals and freelancers (e.g.freelance artisans, tutors and educators, and informal healers or traditional medicine practitioners.). The purposive sampling of the initial participants will be done by leveraging on personal contacts, using the community technical working groups (CTWGs) which is composed of community leaders and representatives of different occupation groups and community health volunteers, and relying on professional networks. Once these participants are identified, they will be consented and recruited for the study. Participants will be requested to refer others from their network who fall under any of the informal sector subgroups. These potential participants in the first wave of snowballing who will be identified, consented and recruited will also be asked for further referrals. Potential participants’ recruitments will be halted when referral chains start overlapping or no new information emerges, or we reach data saturation. Additionally, to enhance the diversity of the sample, participants will be encouraged to refer potential participants from within the same subgroup. To be eligible for inclusion in this study, study participants must be at least 18 years of age.

The districts to be sampled is based on the CHORUS Ghana study sites-Ashaiman and Madina districts of the Greater Accra Region of Ghana.

#### 2.3.3 Step 3: Data Collection-Q-sorting

Participant recruitment and data collection started on the April 1, 2025 and expected to be completed by the 31^st^ of July 2025. In an individual face-to-face interview setting, participants will be required to rank the sampled statements in the Q-set. Participants will be required at this point to rank the sampled statements in the Q-set. This step involves two phases. First, the participants are provided with clear questions probing further on each of the research objectives and then required to rank the sampled statements under each question based on their own preferences on a forced normal distribution curve on a range from most agree (−3) to most disagree (+3). Forced normal distribution will be used to help participants probe deeper into their preferences (46,47). With a forced normal distribution, study participants will be required to rank the sampled statements into a predetermined pattern resembling a normal distribution. This is typically used in q-sort methodology to ensure participants ranking are rated neutrally and fewer items are rated at extreme ends of the scale (46,47).

During the administration of the q-sort, we will conduct, concurrently, interviews with informal sector workers to understand the rationale for their preferred health insurance design features, contextual factors affecting their engagement in health purchasing decisions, and to provide any examples from their experiences with accessing and paying for health care that may have influenced their choices. The q-sorts and interviews will be conducted at the individual level. About 60 interviews will be conducted with informal sector sectors. The demographics of the study participants will be collected. To facilitate the concurrent administration of both the q-sorts and interview, the q-sorts and interviews will be audio recorded.

#### 2.3.4 Step 4: Analysis and Interpretation

##### Quantitative data from Q-sorting

Data analysis will be conducted using the PQMethod software (53). The analysis of q-sort involve correlation, factor analysis and the calculation of factor score (43–45,54). The following steps will be taken to analyse the Q-sorts:

In the first step, the sampled statements would be entered into the PQMethod Software. Then, the data (q-sorts) and demographic data of the study participants will be uploaded into the PQMethod Software.

The next three steps will involve the identification and extraction of factors. Factors refer to a group of persons who have rank ordered the sampled statements in a very similar fashion or a group of people with a similar perspective or viewpoint (55). In step two, a correlation analysis will be conducted. This step is aimed at finding repeated patterns where study participants ranked the statements in identical or similar orders. A correlation matrix will be created by the intercorrelation of each Q-sort with every other sort using Pearson’s R. The correlation matrix shows the nature and extent of the relationship between two Q-sorts and hence the extent of agreement and disagreement in perspectives between study participants (42,55). This helps identify study participants with similar perspectives, that is, similar Q-sorts imply shared perspective. As typical with correlation coefficients, a high correlation coefficient up to 1 suggests a strong association and a lower correlation coefficient suggests a weaker association. Additionally, a positive value of the correlation coefficient suggests a positive relationship whereas a negative correlation coefficient suggests a negative association. For instance, if between participant A and B the correlation coefficient, r, is –0.82, it suggests a high level of disagreement, that is, the statements which A rated highly, are statements B rated poorly and vice versa.

The third step will focus on extracting the initial factors. Once the correlation matrix is created, a factor analysis will be conducted to reduce the data to a smaller number of factors. This is mainly conducted by grouping, comparing and summarising the q-sorts. Study participants with identical viewpoints will have a shared factor. A factor loading is determined for each q-sort, showing the extent to which each q-sort is associated with each factor (43). In this analysis, a Principal Component Analysis (PCA) will be applied. A PCA is a multivariable statistical data reduction technique for minimising the dimensions of analysis. Thus, PCA is a “linear transformation that converts data to a new coordinate system where the greatest variance of any projection of data lies on the first coordinate (the first principal component), the second greatest variance on the second coordinate, and so forth” (56). The PCA will facilitate the transformation of large variables into smaller variables, while retaining a predominant part of information from the original data (57). Another data reducing method that is usually used is the centroid factor analysis. The centroid factor analysis is typically used when there exist a priori assumptions, and researchers extract factors based on theoretical significance or considerations and not objectivity (58). The PCA is preferred over the centroid factor analysis because there were no priori assumptions and preferred the data to lead the way. Besides, as argued by McKeown and Thomas (1988), the difference between the two data reducing methods are negligible (59). The principal strength of PCA over the centroid factor analysis is that PCA provides eigenvalues for respective factors which informs the number of groups you want to build (49). Eigenvalues assess the relative contribution of a factor to the explanation of the total variance in the correlation matrix. Kaiser-Guttman recommends that factors with eigenvalues above 1.00 should be extracted (60). This is because factors with an eigenvalue higher than one explain more variance than a single variable would. Therefore, this study will extract initial or latent variables for the next step of analysis only if their eigenvalues are greater than one.

At the fourth stage, the factors identified (emerging factors) from step three are rotated to derive the final set of factors. The factors are rotated to emphasise the connection between the views of each sorter (43). There are two choices for factor rotation; varimax and hand rotation (54). The varimax procedure allows the researcher to produce a statistical criteria, maximising the eigenvalue for each factor and therefore accounting for the maximum amount of study variance (42,54). This approach is thus independent of the research. Conversely, the hand approach is focused on subjectivity where the researcher positions each factor relative to the other based on a priori theory, or knowledge or substantive knowledge of the q-sort data (Watts and Stenner, 2012). A key strength of the hand approach is that it helps researchers incorporate additional knowledge to the analysis to ensure the factors are rotated to capture and reflect important viewpoints, unlike the varimax approach which is focussed primary on accounting for as much common variance, predominant viewpoints that are exact in the group as a whole (55). However, one predominant setback of the hand approach is the possibility that a factor solution described does not reflect a particular situation but rather reflects the research’s own understanding of the situation (55). This study uses a varimax approach as it is the most efficient way of discovering a subject matter that almost every study participant might recognize and consider to be of importance (55). The final output of a varimax rotation is a matrix of factor loadings, where the study participant’s q-sorts are in rows and the rotated factors as columns (61). The varimax rotation matrix represents the relationship between each q-sort and each rotated factor. Each factor loadings are attached to respective correlation coefficients who show the extent of q sort correlation with each factor (from –1 to +1) (61).

Following factor rotation, the most representative q-sorts for each factor are identified and flagged. Flagging can be conducted either manually based on key assumptions or automatically. In this study, the loading will be significant at p<0.05 level. Thus, all q-sorts that will load significantly on a “single factor and exceed this value either “closely approximated, exemplify, or define the viewpoint of a particular factor””. Only the q-sorts that are flagged will be used in step 6.

The next step involves the calculation of factor scores and other relevant test scores. A factor score that is an estimate of the factor viewpoint, is a weighted average (z-score) of all the individual q-sorts that are associated with that factor (43,55). Based on the z-scores, statements can be attributed to the original quasi-normal distribution, resulting in the conversion of the z-score for each factor into a single composite factor array (42,55). A factor array is a “single q-sort configured to represent the viewpoint of a particular factor” (55).

At this stage, the Watts and Stenner (2012) approach to factor interpretation will be used. In the first phase of the Watts and Stenner (2012) approach, a crib sheet will be created for each factor. The crib sheet will be composed of each statement item organized into a number of categories depending on the nature of the data. The categories mainly consist of the ranking of the statements under each factor which helps you engage with every item. Each factor will be explained in detail with supporting evidence from the qualitative interviews, and demographic data.

##### Qualitative Interviews

The qualitative interviews will be recorded and transcribed verbatim. Guided by the objectives of the study, an appropriate theoretical framework and the themes of the discussion, a coding list will be prepared to guide the data analysis. Thematic analysis will be conducted using QSR NVivo 15 software. The statements of the respondents will be presented as quotes to substantiate the views expressed.

### 2.4 Study Timeline

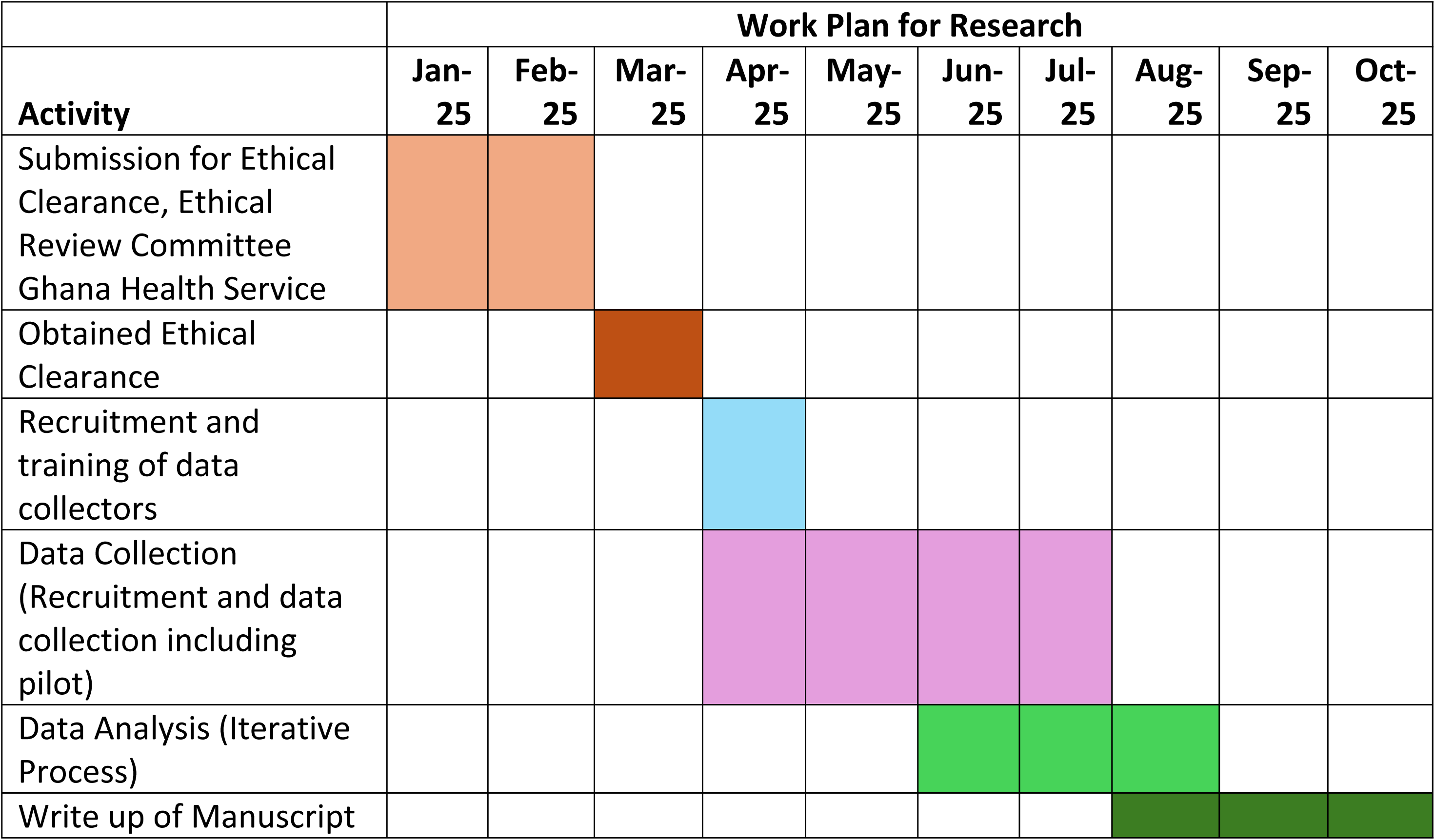

## 3. Discussion

The development of health insurance systems in developing countries such as Ghana are often based on classic formal sector structures although such countries have a predominant informal economy. The findings of this study, which will be presented in webinars and CHORUS consortium meetings, will provide empirical evidence on key design features that are central to improving population coverage rates amongst informal sector workers, strategies to link informal sector workers ability-to-pay to income-based premiums and improve the operational and financial sustainability of health insurance systems. CHORUS partner countries will be at the intervention development stage for subsequent pilots by the time this project is completed. This evidence can easily be factored into these interventions to enhance the responsiveness of system-wide interventions to informal sector workers. Additionally, this evidence would serve as a foundation for advanced studies focusing on how to effectively engage a cadre of stakeholders in health purchasing decisions. It would also serve as a guide to other economic sectors on how to identify informal sector workers and their ability-to-pay for economically viable projects.

## 4. Ethical Consideration

Ethical approval has been obtained from the University of York, Health Science Research Governance Committee (HSRGC/2024/656/C), and the Ghana Health Service Ethics Review Committee (GHS-ERC: 004/02/25). To maintain research integrity, validity and reliability, the steps outlined below will be adhered to. Additionally, research findings will be reported accurately, and any work used that belongs to another author or research will be acknowledged.

**1**. All research participants will be duly consented
  ● Provide research participants with a participant information sheet in a form which can easily be understood. The participant information sheet will include an outline of the objectives of the research study, a summary of the research project, an outline of the responsibility of the research participant and the research investigator, the potential risks and benefits of the research and data accessibility. The participant information sheet will also include the names of all investigators and the details (name, institution affiliated to and contacts) of the principal investigator (PI). This is to facilitate communication between the research participants and the PI, in case research participants have any grievances to report for redressal, or to obtain additional information about the research project. The participant information sheet will also include a statement explaining that participation is entirely voluntary and potential participants are free to decline to participate or have the right to withdraw from the study at any point even after you have consented to participate.
  ● The participant information sheet will be translated according to the language the research participants is most proficient in or comfortable with, if the research participant cannot read or prefers not to read the participant information sheet.
  ● Provide research participants with a consent form. This is a way to document the informed consent process. Each participant will be required to thumbprint or append their signature as proof they have been duly consented.
  ● For the qualitative component of the study, study participants will be consented before they are audio recorded.
**2**. Ensure research participants anonymity and confidentiality
  ● Research participants will be administered anonymous questionnaires. No questionnaire will include any identifiers and any identifier that is found will be removed during the data processing and analysis. Pseudonym names will be used particularly for the qualitative data
  ● For the qualitative research, identified key stakeholders or stakeholders may be requested to help identify potential research participants. Thus, the anonymity of some research participants may not be guaranteed. To resolve this, I will request for a list of potential participants and select a few from the list.
  ● Although I will collate the contact details of potential research participants (key stakeholders) to arrange for qualitative interviews, such information will be stored securely and discarded after research dissemination.
  ● All interviews will be conducted at a private environment that is conducive and approved by the research participant
  ● All endorsed consent forms will be secured in a locked cabinet. I will be the only who would have access to the consent forms until time to discard consent forms per the regulations of the University of York
  ● Transcribers will be duly informed of the ethics of research and the need to ensure research participants anonymity and confidentiality to prevent them from discussing the audio interviews
  ● All data will be secured, analyzed and findings disseminated in accordance with the University of Hull Data Protection Policies
  ● University of York online cloud storage OneDrive will be used to securely store audio recordings, transcripts, and data from the surveys. The University of York Google Drive will be used as a backup system.
  ● The quantitative data will be stored in a PQMethod format. Analysis will thus be conducted in the PQMethod Software. The qualitative data will be transcribed verbatim and exported to NVivo for data analysis.
  ● Data will be shared with investigators on the CHORUS Ghana project, the main project this PhD research is linked to, under a data sharing agreement.
  ● The findings of the study will be disseminated through peer-reviewed journals, seminars, symposiums and relevant health systems or financing meetings.
**3**. Ensure beneficence, non-maleficence, and justice

● All research participants will be treated with respect and dignity during the study
● Research participants for the focus group discussions will be systematically selected to ensure fair representation of all relevant groups particularly groups that will directly benefit from the research
● The focus group discussions will also be organized in a manner that ensures participants express their views without fear or malice.
● All individuals who will be included in this study will be referred to as study participants or stakeholders. No individual will be referred to or regarded as a subject.

## 5. Dissemination

The findings of this research supplements existing efforts to improve engagement between communities, key stakeholders within the health and local government systems to improve primary health outcomes. Empirical evidence from this research will reinforce advocacy for and the formulation of policies specific to improving informal sector workers uptake and use of health insurance. The findings of the study would be disseminated through two main publications; a peer-reviewed journal article and a policy brief. Additionally, the findings of the study would be presented at appropriate conferences, domestic and international, and annual health review meetings, and CHORUS consortium meetings.

## 6. Limitations

This study is subject to a number of limitations. First, the use of the Q-sort methodology while effective for exploring subjective viewpoints, involves a relatively small, purposively selected sample. This limits the generalizability of findings to all informal sector workers in Ghana or other LMICs. Additionally, purposive and snowball sampling may introduce selection bias, potentially excluding more marginalized individuals within the informal sector. Second, while efforts will be made to ensure content and face validity of the Q-set statements, participants may interpret statements differently based on personal experiences, particularly given the complex nature of health insurance design. This subjectivity could affect the consistency and comparability of Q-sort responses. Third, despite efforts to minimize language and literacy barriers, these barriers may affect some participants’ ability to understand and accurately respond to Q-sort statements, even with translated materials and piloting.

## 7. Conclusion

This research will provide new knowledge on health insurance designs which reflect the preferences, values and needs of informal sector workers and health facilities in Ghana. It will particularly provide valuable insights on how to effectively engage a cadre of stakeholders in health purchasing decisions to enhance enrollment of informal sector workers unto the Ghana NHIS.

## Acknowledgements

We acknowledge Max Nawrath’s critical review of the study protocol and for providing constructive guidance on the data collection approach.

## Funding

This study is part of a PhD Research which is funded by the Hull and York Medical School, University of York

## Competing Interests

No authors have competing interests.

## Data Availability

Because this article follows a protocol format, no datasets were created or analyzed in its preparation.

## Supporting Information

**S1 Appendix: In-Depth Interviews with Informal Sector Workers**

**S2 Appendix: In-Depth Interviews with Health Providers**

**S3 Appendix: In-Depth Interviews with Key Health and Policy Stakeholders**

## Notes

### Competing Interest Statement

The authors have declared no competing interest.

### Funding Statement

Yes

### Author Declarations

Ethical approval was obtained from the University of York, Health Science Research Governance Committee (HSRGC/2024/656/C), and the Ghana Health Service Ethics Review Committee (GHS-ERC: 004/02/25).

